# Predicting Functional Dependency in Patients with Disorders of Consciousness: A TBI-Model Systems and TRACK-TBI Study

**DOI:** 10.1101/2023.03.14.23287249

**Authors:** Samuel B. Snider, Nancy R. Temkin, Jason Barber, Brian L. Edlow, Joseph T. Giacino, Flora M. Hammond, Saef Izzy, Robert G. Kowalski, Amy J. Markowitz, Craig A. Rovito, Shirley L. Shih, Ross D. Zafonte, Geoffrey T. Manley, Yelena G. Bodien, The TRACK-TBI Investigators

**Affiliations:** Division of Neurocritical Care, Department of Neurology, Brigham and Women’s Hospital, Boston, MA, USA; Harvard Medical School, Boston, MA, USA; Department of Neurological Surgery, University of Washington, Seattle, Washington, USA; Department of Biostatistics, University of Washington, Seattle, Washington, USA; Center for Neurotechnology and Neurorecovery and Department of Neurology, Massachusetts General Hospital, Boston, MA, USA; Athinoula A. Martinos Center for Biomedical Imaging, Massachusetts General Hospital, Charlestown, MA, USA; Department of Physical Medicine and Rehabilitation, Spaulding Rehabilitation Hospital, Boston, MA USA; Department of Physical Medicine and Rehabilitation, Indiana University School of Medicine, Indianapolis, IN, USA; Departments of Neurosurgery and Neurology, University of Colorado School of Medicine, Aurora CO, USA; Department of Neurological Surgery, UCSF, San Francisco, CA USA; Brain and Spinal Cord Injury Center, Zuckerberg San Francisco General Hospital and Trauma Center, San Francisco, CA USA; University of Maryland; MassGeneral Hospital for Children; University of Cincinnati; Baylor College of Medicine; University of Utah; University of Washington; Medical College of Wisconsin; Virginia Commonwealth University; University of California, San Francisco; University of Pittsburgh; UT Austin

**Keywords:** Traumatic brain injury, Disorders of Consciousness, Dependency, Outcomes, Prediction

## Abstract

**Importance:** There are currently no models that predict long-term functional dependency in patients with disorders of consciousness (DoC) after traumatic brain injury (TBI).

**Objective:** Fit, test, and externally validate a prediction model for 1-year dependency in patients with DoC 2 or more weeks after TBI.

**Design:** Secondary analysis of patients enrolled in TBI Model Systems (TBI-MS, 1988-2020, Discovery Sample) or Transforming Research and Clinical Knowledge in TBI (TRACK-TBI, 2013-2018, Validation Sample) and followed 1-year post-injury.

**Setting:** Multi-center study at USA rehabilitation hospitals (TBI-MS) and acute care hospitals (TRACK-TBI).

**Participants:** Adults with TBI who were not following commands at rehabilitation admission (TBI-MS; days post-injury vary) or 2-weeks post-injury (TRACK-TBI).

**Exposures:** In the TBI-MS database (model fitting and testing), we screened demographic, radiological, clinical variables, and Disability Rating Scale (DRS) item scores for association with the primary outcome.

**Main Outcome:** The primary outcome was death or complete functional dependency at 1-year post-injury, defined using a DRS-based binary measure (DRS_Depend_), indicating need for assistance with all activities and concomitant cognitive impairment.

**Results:** In the TBI-MS Discovery Sample, 1,960 subjects (mean age 40 [18] years, 76% male, 68% white) met inclusion criteria and 406 (27%) were dependent at 1-year post-injury. A dependency prediction model had an area under the receiver operating characteristic curve (AUROC) of 0.79 [0.74, 0.85], positive predictive value of 53%, and negative predictive value of 86% for dependency in a held-out TBI-MS Testing cohort. Within the TRACK-TBI external validation sample (N=124, age 40 [16], 77% male, 81% white), a model modified to remove variables not collected in TRACK-TBI, had an AUROC of 0.66 [0.53, 0.79], equivalent to the gold-standard IMPACT_core+CT_ score (0.68; 95% AUROC difference CI: -0.2 to 0.2, p=0.8).

**Conclusions and Relevance:** We used the largest existing cohort of patients with DoC after TBI to develop, test and externally validate a prediction model of 1-year dependency. The model’s sensitivity and negative predictive value were greater than specificity and positive predictive value. Accuracy was diminished in an external sample, but equivalent to the best-available models. Further research is needed to improve dependency prediction in patients with DoC after TBI.

## Introduction

Most patients with disorders of consciousness (DoC) after traumatic brain injury (TBI) recover consciousness^1-3^. A recent TBI Model Systems (TBI-MS) study demonstrated that among patients admitted to inpatient rehabilitation with DoC, 82% recovered consciousness and 40% recovered some degree of independence by rehabilitation discharge^1^. However, the recovery endpoint can vary widely, from complete dependency (i.e., inability to perform basic self-care) to return to pre-injury level of function^2-6^. Accurately predicting this outcome could reduce the heterogeneity observed in clinical prognostication^7^ and aid in the meaningful stratification of patients for clinical trial enrollment.

Despite its importance to quality of life, health care expenditure, and caregiver burden^8-13^, the predictors of long-term functional dependency in patients with DoC after TBI are uncertain^14^. Though post-acute DoC diagnoses are associated with outcome^6, 15-17^, the clinical characteristics that predict prolonged dependency are not well established. Two models have been validated to predict death or severe disability after TBI^18-20^. However, the accuracy of such models in patients who survive the initial injury with DoC is uncertain.

Assessment of dependency is limited by the available outcome scales, including the Glasgow Outcome Scale Extended [GOSE] ^21, 22^ and the Disability Rating Scale (DRS)^23^, which may lack sensitivity or specificity for capturing dependency in severely-injured patients.^4, 24^ Furthermore, the frequently applied endpoint of 6-months post-injury may be too early, as severely injured patients are likely still recovering^6, 25, 26^. We recently developed a novel measure based on the DRS, the DRS_Depend_, that identified patients with complete dependency 1 year after TBI more accurately than either the GOSE or the total DRS score^4^.

To improve the accuracy of outcome prediction in patients with DoC after TBI, we aimed to: 1) develop and internally test a dependency (DRS_Depend_) prediction model at 1-year post-injury in a large national database (TBI Model Systems [TBI-MS]^27^); 2) evaluate model performance in an independent sample of patients enrolled in the Transforming Research and Clinical Knowledge in TBI (TRACK-TBI)^28^ study; and 3) compare our model’s performance to the International Mission for Prognosis and Analysis of Clinical Trials in TBI (IMPACT)^18^ score, a frequently-used, validated^20^ prognostic model.

## METHODS

We conducted a secondary analysis of two prospective, longitudinal cohorts: TBI-MS National Database^27^ and TRACK-TBI^28^ (clinicaltrials.gov NCT02119182; https://tracktbi.ucsf.edu/). Characteristics of each cohort have been previously described^1, 2, 28-31^. Briefly, TBI-MS enrolls adults (≥ 16 years) with moderate or severe TBI who survive acute hospitalization and are admitted to inpatient rehabilitation centers in the US. 25 centers have contributed data since 1988. TBI-MS participants are followed at 1, 2, 5, and every 5-years post-injury to assess outcomes across a range of domains. TRACK-TBI enrolls participants with TBI and a clinically ordered CT scan presenting to 18 Level 1 trauma centers in the US. TRACK-TBI participants are assessed across a range of domains at 2-weeks, 3-months, 6-months, and 1-year post-injury^32^. Local Institutional Review Boards at each site provide study approval, and participants or their surrogates provide written informed consent. The Mass General Brigham Institutional Review Board provides local oversight and approved this study (IRB Protocols: 2012P002476, 2013P002241).

### Study Samples

We used the TBI-MS database (i.e., Discovery Sample) to fit and internally test the dependency model because of its large sample size (N > 2,000 with DoC at rehabilitation admission), which offers a unique opportunity to identify factors associated with long-term dependency.

The TBI-MS database enrolls only survivors of the acute injury, mitigating the self-fulfilling prophecy bias^33^ resulting from including patients with early death in prediction models. We externally validated our model in a cohort of patients with DoC at 2-weeks post injury enrolled in TRACK-TBI.

#### TBI-MS (Discovery Sample)

TBI-MS participants are enrolled and initially assessed at the time of admission to an inpatient rehabilitation hospital, which immediately follows discharge from acute care^1, 34^. We identified patients with DoC on admission to inpatient rehabilitation using the same criteria applied in prior studies^1, 3^, an admission DRS_Motor_ item score > 0 (i.e., absence of command following). We excluded patients missing DRS scores on admission to rehabilitation (N=218). We also excluded patients with outlier acute care lengths of stay, (N=167; 1.5 x interquartile range [IQR]: > 55 days; Supplementary Figure 1), as the first study assessment in these patients would occur nearly 2 months post injury. Within this Discovery Sample, participants were randomly assigned to 80% Model Derivation and 20% Model Testing cohorts, holding the outcome proportions constant in each.

#### TRACK-TBI (External Validation Sample)

To assess the external validity of our prediction model, we defined an analogous cohort of adults (≥17-years-old) in TRACK-TBI, with moderate or severe TBI (Glasgow Coma Scale [GCS] Score < 13 on arrival to the emergency department), and DoC at the 2-week post-injury standard assessment (not following commands; DRS_Motor_ > 0; Supplementary Figure 2). TBI-MS and TRACK-TBI differ in terms of the minimum TBI severity required for enrollment (moderate [TBI-MS] vs mild [TRACK-TBI]), location of enrollment (inpatient rehabilitation hospital [TBI-MS] versus acute care hospital [TRACK-TBI]), and the timing of first post-injury DRS assessment (admission to rehabilitation, which does not occur at a fixed time-point [TBI-MS] versus 2-weeks post-injury [TRACK-TBI]).

### Primary Outcome

Our primary outcome was complete functional dependency at 1-year post-injury, defined as a positive DRS_Depend_ score (DRS_Depend_+). The DRS_Depend_ is a binary DRS-composite score corresponding to a DRS_Function_ item score ≥ 4 (indicating the need for assistance with all activities at all times), *and* a score of > 0 (some assistance needed) on at least one of the following items: cognitive ability for Communicating, Feeding, Toileting, or Grooming^4^. Participants meeting DRS_Depend_ criteria require the assistance of another person at all times due, at least in part, to cognitive impairment^4^.

In the primary analysis, the DRS_Depend_+ outcome category included participants who died after admission to rehabilitation, but before the 1-year follow-up. In a secondary analysis, we treated these deaths as missing outcome data, given the inherent uncertainty about the ultimate level of recovery in such patients. We considered survivors without a documented DRS at 1-year (365 +/- 62 days post-injury; N=428, Table 1) to have missing outcome data, as recommended by the TBI-MS standard operating procedure^34^.

**Table 1:** TBI-MS Cohort Characteristics

|  | 1-Year Outcome |  |  |  | All<br>(N = 1,960) |
| --- | --- | --- | --- | --- | --- |
|  | DRS<sub>Depend-</sub><br>(N = 1126) | DRS<sub>Depend+</sub><br>(N = 303) | Died<br>(N = 103) | Missing<br>(N = 428) |  |
| DEMOGRAPHICS |  |  |  |  |  |
| Age; Mean (SD) | 38 (18) | 43 (20) | 58 (22) | 41 (19) | 40 (19) |
| Sex Male; N (%) | 866/1125 (77) | 226/301 (75) | 78/103 (76) | 317/428 (74) | 1487/1957 (76) |
| Race White; N (%) | 791/1126 (70) | 193/303 (64) | 77/103 (75) | 275/428 (64) | 1336/1960 (68) |
| Marital status; N (%) |  |  |  |  |  |
| Single | 565/1124 (50) | 151/303 (50) | 31/103 (30) | 195/426 (46) | 942/1956 (48) |
| Married | 361/1124 (32) | 104/303 (23) | 40/103 (39) | 143/426 (33) | 648/1956 (33) |
| Other | 198/1124 (18) | 48/303 (16) | 32/103 (31) | 88/426 (21) | 366/1956 (19) |
| Education; N (%) |  |  |  |  |  |
| < HS | 334/986 (34) | 80/273 (29) | 21/87 (24) | 96/303 (32) | 531/1649 (32) |
| HS diploma/GED | 280/986 (28) | 88/273 (32) | 34/87 (39) | 106/303 (35) | 508/1649 (31) |
| Some college | 178/986 (18) | 45/273 (17) | 16/87 (18) | 44/303 (15) | 283/1649 (17) |
| Batchelor's Degree | 194/986 (20) | 60/273 (22) | 16/87 (18) | 57/303 (19) | 327/1649 (20) |
| Employment; N (%) |  |  |  |  |  |
| Competitively Employed | 622/967 (64) | 153/273 (56) | 25/94 (27) | 166/298 (56) | 966/1632 (59) |
| Student | 76/967 (8) | 13/273 (5) | 3/94 (3) | 21/298 (7) | 113/1632 (7) |
| Unemployed | 269/967 (28) | 107/273 (39) | 66/94 (70) | 111/298 (37) | 553/1632 (34) |
| Housing; N (%) |  |  |  |  |  |
| Private Home | 1108/1126 (98) | 296/302 (98) | 96/102 (94) | 416/427 (97) | 1916/1957 (98) |
| Other | 18/1126 (2) | 6/302 (2) | 6/102 (6) | 11/427 (3) | 41/1957 (2) |
| CLINICAL CHARACTERISTICS |  |  |  |  |  |
| Injury Mechanism; N (%) |  |  |  |  |  |
| High-Velocity | 690/1123 (61) | 149/301(50) | 29/103 (28) | 198/427 (46) | 1066/1954 (55) |
| Fall-related | 236/1123 (21) | 89/301 (30) | 54/103 (52) | 103/427 (24) | 482/1954 (25) |
| Low-Velocity/Other | 197/1123 (18) | 63/301 (21) | 20/103 (19) | 126/427 (30) | 406/1954 (21) |
| Injury Year; Mean (SD) | 2010 (6) | 2010 (6) | 2010 (6) | 2010 (8) | 2010 (7) |
| ED GCS<sub>Total</sub>; Median (IQR) | 7 (9) | 8 (9) | 12 (9) | 9 (9) | 8 (9) |
| ED GCS<sub>Verbal</sub> < 3; N (%)<sup>a</sup> | 315/1123 (28) | 73/299 (24) | 28/103 (27) | 98/427 (23) | 514/1952 (26) |
| ED GCS<sub>Motor</sub> < 5; N (%)<sup>b</sup> | 513/1123 (46) | 149/299 (49) | 35/103 (34) | 196/427 (46) | 893/1952 (46) |
| ED GCS<sub>Eye</sub> < 2; N (%)<sup>b</sup> | 505/1123 (45) | 137/299 (45) | 33/103 (32) | 198/427 (46) | 873/1952 (45) |
| Intubated; N (%) | 606/1126 (54) | 177/303 (58) | 41/103 (39) | 240/428 (56) | 1064/1960 (54) |
| Craniectomy; N (%) | 138/906 (15) | 75/262 (29) | 16/90 (18) | 54/265 (20) | 283/1523 (19) |
| <u>CT Compression; N (%)</u> |  |  |  |  |  |
| None | 536/923 (58) | 115/251 (46) | 40/91 (44) | 180/314 (57) | 871/1579 (55) |
| Cisterns present/MLS<5mm | 89/923 (10) | 30/251 (12) | 12/91 (13) | 29/314 (9) | 160/1579 (10) |
| Cisterns absent/MLS>5mm | 298/923 (32) | 106/251 (42) | 39/91 (43) | 105/314 (33) | 548/1579 (35) |
| Multiple Episodes of Intracranial Hypertension; N (%) | 281/1112 (25) | 96/294(32) | 18/102 (18) | 85/422 (20) | 480/1930 (25) |
| SDH/SAH (%) | 914/1077 (85) | 262/283 (93) | 88/97 (91) | 300/367 (82) | 1564/1824 (86) |
| EDH | 117/1073 (11) | 35/282 (12) | 11/98 (11) | 39/367 (11) | 202/2830 (11) |
| IVH | 406/1076 (38) | 154/283 (54) | 40/98 (41) | 128/367 (35) | 728/1824 (40) |
| <u>Contusions</u> |  |  |  |  |  |
| Any; N (%) | 812/1076 (76) | 222/283 (78) | 71/98 (72) | 283/367 (77) | 1388/1824 (76) |
| Frontal; N (%) | 665/1076 (62) | 184/283 (65) | 60/98 (61) | 229/367 (62) | 1138/1824 (62) |
| Temporal; N (%) | 498/1076 (46) | 144/282 (51) | 39/98 (40) | 170/366 (46) | 851/1822 (47) |
| Parietal; N (%) | 235/1076 (22) | 76/283 (27) | 18/98 (18) | 83/367 (23) | 412/1824 (23) |
| Occipital; N (%) | 91/1076 (9) | 32/282 (11) | 3/98 (3) | 34/367 (9) | 160/1823 (9) |
| Non-cortical; N (%) | 323/1075 (30) | 121/282 (43) | 22/98 (22) | 104/367 (28) | 570/1822 (31) |
| Total #; Mean (SD) | 2 (2) | 3 (2) | 2 (2) | 2 (2) | 2 (2) |
| Early command following [within 5 days of injury]; N (%) | 288/1121 (26) | 37/302 (12) | 36/103 (35) | 102/424 (24) | 463/1950 (24) |
| Days post-injury [*first assessment*]*; Mean (SD) | 24 (11) | 30 (12) | 24 (13) | 24 (12) | 25 (12) |
| DRS<sub>Depend</sub>+ [*first assessment*]*; N(%) | 1058/1124 (94) | 295/299 (99) | 98/102 (96) | 403/425 (95) | 1854/1950 (95) |
| <u>DRS [*first assessment*]; Median(IQR)</u> |  |  |  |  |  |
| Total Score | 21 (5) | 23 (4) | 22 (4) | 21 (5) | 22 (4) |
| DRS<sub>Function</sub> | 5 (1) | 5 (0) | 5 (0) | 5 (0) | 5 (0) |
| DRS<sub>Eye</sub> | 0 (1) | 1 (2) | 1 (2) | 0 (1) | 0 (1) |
| DRS<sub>Motor</sub> | 1 (1) | 2 (2) | 2 (2) | 1 (1) | 1 (1) |
| Rehabilitation LOS; Mean (SD) | 44 (34) | 74 (74) | 40 (30) | 51 (43) | 50 (45) |
| **OUTCOMES** |  |  |  |  |  |
| DRS [*1-year*]; Median (IQR) | 3 (3) | 14 (5) | 30 (0) | - | 7 (8) |
| GOSE [*1-year*]; Median (IQR) | 5 (2) | 3 (1) | 1 (0) | 4 (1) | 4 (2) |

a = in patient who is not intubated
b = in patient not receiving sedation/paralytic
\* First assessment at rehabilitation admission
Abbreviations: ED = Emergency Department; admit = admission; SD = Standard Deviation; IQR = Interquartile Range; GED = Geeral Educational Development Test; MLS = Midline Shift; SDH = Subdural Hemorrhage; SAH = Subarachnoid Hemorrhage; EDH = Epidural Hemorrhage; IVH = Intraventricular Hemorrhage; DRS = Disability Rating Scale; GOSE = Glasgow Outcome Scale Extended; LOS = Length of Stay

To mitigate potential bias from missing outcome data, we used inverse propensity weighting with gradient boosted models (R: TWANG package), separately in each dataset (TBI-MS and TRACK-TBI), to assign participants with complete outcome data a weight based on their clinical and demographic similarity to subjects with missing outcome data (Supplementary Figure 3). We used default settings, truncated the weights at the 99^th^ percentile, normalized by the sample mean, and applied the scaled weights to the subsequent logistic regressions.

### Exposure Variables

Within the TBI-MS Discovery Sample, we screened demographic, clinical, and radiological variables (e.g., presence of contusion in a specific brain lobe) for a possible association with the primary outcome. All variables were prospectively collected according to standardized criteria, as documented in the TBI-MS standard operating procedure^34^. A complete list of variables and their definitions is provided in the Supplementary Material. Among variable pairs with > 70% collinearity by Spearman’s Rho, only one variable was analyzed further (Supplementary Figure 4). Continuous and ordinal variables were factorized based on their observed univariate association with the primary outcome in the 80% TBI-MS Model Derivation cohort.

We excluded variables with missing entries in more than 30% of participants (Supplementary Figure 5). Otherwise, missing exposure variables were imputed using multiple imputation with chained equations (R software: MICE package). The imputation model was estimated in only the 80% training sample, with weights subsequently applied to the 20% testing sample. As previously recommended,^35^ we imputed 5 datasets with 50 iterations per imputation.

### Statistical Approach

#### TBI-MS (Discovery Sample)

Within the TBI-MS 80% Model Derivation cohort, we performed a univariate logistic regression screen of all exposure variables, retaining variables with p < 0.1 (pooled estimate across imputed datasets) for association with the outcome. To identify a minimal set of variables with the best outcome-explanatory power while avoiding overfitting, we used Bayesian Information Criterion (BIC)^36^ based reverse selection. BIC-based selection is slightly more restrictive (favoring fewer variables in the model) than Akaike Information Criterion-based selection^37^. We identified variables selected in at least 4 of 5 imputed datasets and fit a final model in each imputed dataset with these variables, pooling the final coefficient estimates and standard errors across datasets.

We evaluated model performance using Area Under the Receiver Operating Characteristic (AUROC) in the 20% TBI-MS Model Testing cohort. We generated model predictions across each imputed dataset and averaged them in accordance with Rubin’s Rules for prediction using multiply-imputed datasets^35, 38^. We defined the optimal threshold for predicting dependency as the point that maximized the Youden J Statistic (sensitivity+specificity-1). Given our intention to create a generalizable model, we then excluded TBI-MS variables not collected in the TRACK-TBI study or with the potential to reinforce existing sociodemographic disparities and fit a second “TRACK-TBI aligned” model using the procedure described above.

#### External Validation

To assess external validity of the model, we applied the “TRACK-TBI aligned” prediction model to patients meeting inclusion criteria in the TRACK-TBI dataset. We tested whether the discrimination performance of our model was different from chance and from the IMPACT^18^ prediction model (Delong’s Test).

## RESULTS

### TBI-MS (Discovery Sample)

Among 18,486 subjects in the TBI-MS database enrolled between 10/1/1988 and 09/04/2020, 2,127 (12%) met criteria for DoC (i.e., not following commands) at first assessment (rehabilitation admission). Our final cohort (TBI-MS Discovery Sample) included 1,960 participants (Figure 1). At 1-year post-injury, 428 (22%) were lost to follow-up or had missing DRS assessment, and the primary outcome (death or DRS_Depend_+) occurred in 406 (27%).

**Figure 1:**
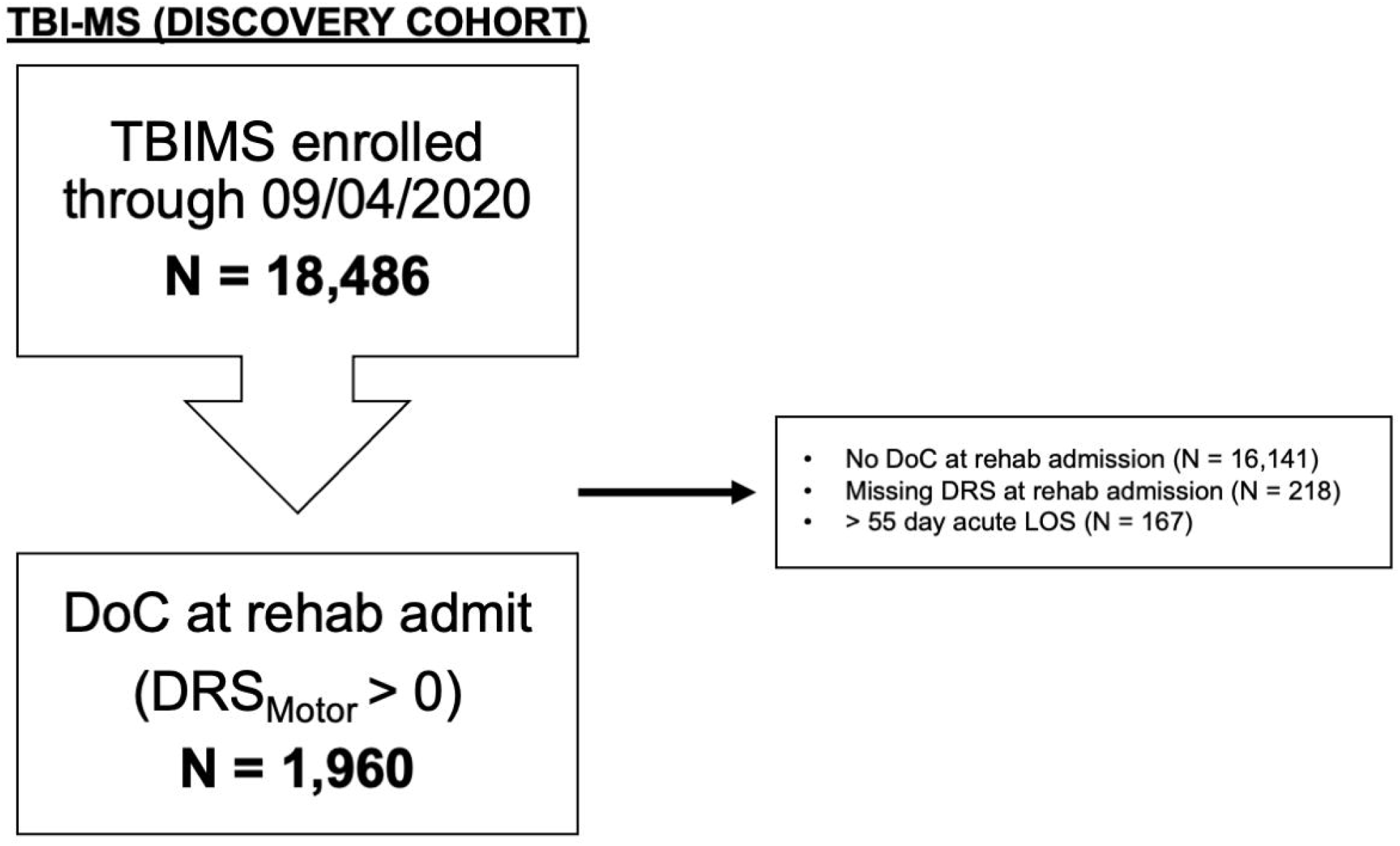
TBI-MS Cohort Flowchart. TBI-MS (Discovery Cohort) CONSORT flow diagram. Abbreviations: DOC = Disorder of Consciousness; DRS = Disability Rating Scale, DRSm = DRS_Motor_ sub score

Participants included in the Discovery Sample were mean (SD) 40 (±19) years old, predominantly white (N=1,336 [68%]) and male (N=1,487 [76%], Table 1). A high-velocity mechanism of injury was reported in 1,066 (55%) and the median [Interquartile range, IQR] GCS on arrival to the acute care hospital was 8 [4, 13]. Our inclusion criteria stipulated that participants did not follow commands on DRS assessment at rehabilitation admission. However, based on clinical notes, 463 (24%) followed commands at least once within 5 days of injury, suggesting a fluctuating neurological examination. Contusions were documented on initial CT imaging in 1,388 (76%) and intraventricular hemorrhage (IVH) in 728 (40%). At first assessment (mean 25±12 days post-injury), 1,854 (95%) were DRS_Depend_+, and the median [IQR] DRS total score was 22 [19, 23] (vegetative state)^2^.

### Deriving and Testing TBI-MS Prediction Model (Discovery Sample)

The randomly partitioned Model Derivation (i.e., 80%, N=1,226) and Model Testing (i.e., 20%, N=306) cohorts were well-matched (see: Supplementary Table 1 for cohort differences). All variables with a univariate association (p≤0.1) with the primary outcome in the Model Derivation cohort are shown in Supplementary Table 2.

**Table 2:** Comparison of Model Testing (TBI-MS) and External Validation (TRACK-TBI) Cohorts

|  | **TBI-MS Testing Cohort**<br>**(N = 306)** | **TRACK-TBI External Validation Cohort**<br>**(N = 124)** | **P value** |
| --- | --- | --- | --- |
| **DEMOGRAPHICS** |  |  |  |
| Age; Mean (SD) | 42 (20) | 40 (16) | 0.4 |
| Sex Male; N (%) | 224/306 (73) | 95/124 (77) | 0.5 |
| Race White; N (%) | 197/306 (64) | 100/123 (81) | 0.001 |
| Education Years; Mean (SD) | 12 (3) | 12 (2) | 0.7 |
| **CLINICAL CHARACTERISTICS** |  |  |  |
| Craniectomy; N (%) | 49/259 (19) | 52/122 (43) | < 0.0001 |
| IVH; N (%) | 127/294 (43) | 28/116 (24) | 0.0005 |
| <u>DRS<sub>Function</sub> [first assessment]*; N (%)</u> |  |  |  |
| Not Totally-dependent [score=1-4] | 72/305 (24) | 2/124 (2) | < 0.0001 |
| Totally-dependent [5] | 233/305 (76) | 122/124 (98) |  |
| <u>DRS<sub>Eye</sub> [first assessment]*; N (%)</u> |  |  |  |
| Opens eyes spont/to voice [0-1] | 256/306 (84) | 35/123 (28) | < 0.0001 |
| Opens eyes to nox stim [2] | 32/306 (10) | 32/123 (26) |  |
| Does not open eyes [3] | 18/306 (6) | 56/123 (45) |  |
| <u>DRS<sub>Motor</sub> [first assessment]*; N (%)</u> |  |  |  |
| Localizing [1] | 188/306 (61) | 13/123 (10) | < 0.0001 |
| Withdrawal [2] | 68/306 (22) | 34/123 (28) |  |
| Flex/extend/no mvmt [3-5] | 50/306 (16) | 76/123 (62) |  |
| Days post-injury [first assessment]*; Mean (SD) | 24 (11) | 14 (3) | < 0.0001 |
| Followed cmds first 5 days; N (%) | 68/305 (22) | 20/101 (20) | 0.7 |
| Discharged to inpatient rehabilitation; N (%) | 306/306 (100) | 45/115 (39) | < 0.0001 |
| **OUTCOMES** |  |  |  |
| DRS<sub>Depend</sub> + [1-year]; N (%) | 59/306 (19) | 35/100 (35) | 0.002 |
| Died; N (%) | 22/306 (7) | 22/100 (22) | < 0.0001 |
| GOSE 2-3 [1-year]; N (%) | 114/287 (40) | 39/100 (39) | 0.8 |
| Missing Outcome; N (%) | -- | 24/100 (24) |  |

\* First assessment at rehabilitation admission (TBI-MS) and 2 weeks post injury (TRACK)
**Abbreviations:** SD = standard deviation; IVH = intraventricular hemorrhage; nox = noxious; DRS = Disability Rating Scale; spont = spontaneous; cmds = commands; GOSE = Glasgow Outcome Scale Extended (GOSE)

The final model is shown in Figure 2A and Supplementary Table 3. The following variables were associated with dependency: age > 55 (adjusted Odds Ratio [aOR]: 4.3 [3.0, 6.1]), first assessment DRS_Function_ sub-scale score (*Totally Dependent:* 2.6 [1.7, 4.0]), first assessment DRS_Motor_ sub-scale score (*Flexion/extension/no-movement*: 3.4 [2.4, 4.9]; *Withdrawal from noxious stimulation*: 1.7 [1.2, 2.4]), craniectomy (1.8 [1.3, 2.5]), IVH on initial CT (1.6 [1.2, 2.1]), non-cortical contusion on initial CT (1.7 [1.2, 2.4]), followed commands within 5 days of injury (0.5 [0.3, 0.7]) and pre-injury employment status (*Unemployed*: 2.1 [1.5, 2.9]; *student*: 0.7 [0.4, 1.2]).

**Figure 2:**
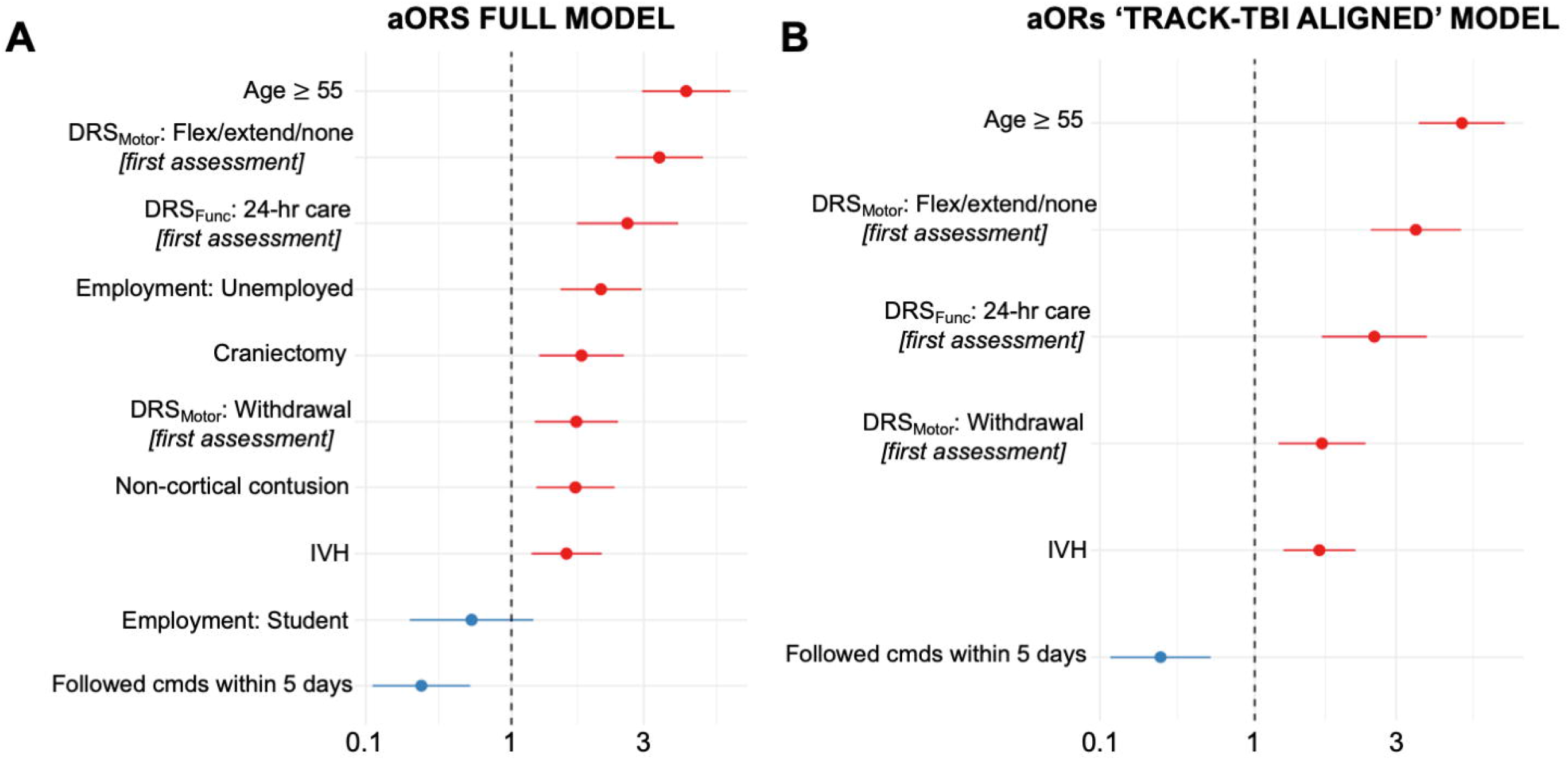
Dependency Prediction Models. (A) A forest plot of adjusted odds ratios (aORs) for variables included in the dependency prediction model, with deaths during follow-up included as dependency. (B) A forest plot of variables included in the “TRACK-TBI Aligned” model. DRS: Disability Rating Scale, Cmds: commands, IVH: intraventricular hemorrhage

When we tested model performance within the separate TBI-MS Model Testing cohort, our prediction model had an AUROC of 0.79 [0.74, 0.85] (Figure 3A). Using the Youden-optimal probability threshold of 0.34 (Figure 3B), the model identified dependency with 65% sensitivity and 79% specificity, positive predictive value (PPV) of 53% and negative predictive value (NPV) of 86%. Model predictions were well-calibrated (Figure 3C; Hosmer and Lemeshow goodness of fit test: *X*^2^_(8,306)_=7.5, p=0.5).

**Figure 3:**
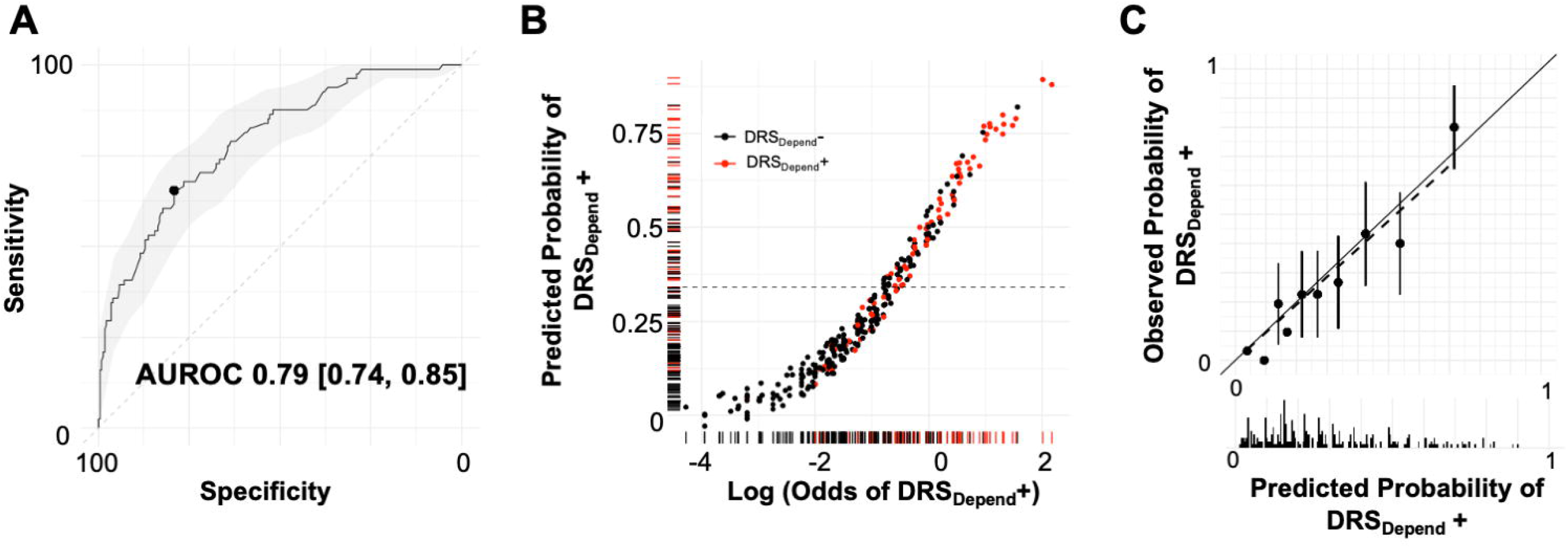
Performance in TBI-MS Model Testing Sample. (A) Area under the Receiver Operating Characteristic Curve for TBI-MS Dependency prediction model within the TBI-MS Testing Sample. Shaded area reflects the 95% confidence interval. (B) Rug plot showing predicted dependency probabilities (y axis) and the logarithm of the odds of dependency (x axis). Each dot represents one subject and each subject is also entered as a tick on each axis. Color indicates 1-year outcome (red: DRS_Depend_+, black: DRS_Depend_ -). The dashed line represents the Youden J cut-point (0.34) derived from (A). DRS_Depend_+ subjects cluster towards the upper right quadrant of the plot. (C) Calibration plot showing predicted dependency probabilities (x axis) and observed probabilities (y axis). Each dot represents a 10-percentile bin of predicted probabilities. Error bars represent 95% confidence intervals. Histogram below the x axis shows the relative density of observations in each predicted probability bucket.

We subsequently developed a “TRACK-TBI Aligned” model, excluding variables not collected in the TRACK-TBI study (e.g., non-cortical contusion) and potentially bias-reinforcing sociodemographic variables (e.g., employment status). This model (Figure 2B; Supplementary Table 4) had undiminished performance in the TBI-MS Testing cohort (Supplementary Figure 6; optimal cut-point 0.28, sensitivity 75%, specificity 72%, PPV 50%, NPV 89%, and AUROC of 0.78 [0.72, 0.84]; 95% CI for AUROC difference between models: -0.04 to 0.02, p = 0.6; Hosmer and Lemeshow: *X*^2^_(8,306)_=7.8, p=0.5).]

We observed that the association between rehabilitation admission DRS_Motor_ score and 1-year dependency varied by time (post-injury) to rehabilitation admission (aOR 1.04 [1.01, 1.08] per additional day; p=0.01; Supplementary Figure 7). We did not include this interaction in the model because in the TBI-MS dataset, time post-injury also encodes acute care length of stay, a variable that would not be available when applying the model at a fixed post-injury time point. To ensure our results were not dependent on the inclusion of deaths in our primary outcome, we repeated our analysis treating deaths as missing outcomes, which resulted in a model that was nearly identical to our original model (Supplementary Figure 8).

### External Validation of the TRACK-TBI-Aligned Model

Among N = 2,552 participants enrolled in the TRACK-TBI, 124 (5%) met inclusion criteria at 2-weeks post-injury (Supplementary Figure 2). Twenty-four (19%) were lost to follow-up and 57 (57%) met criteria for the primary outcome (dead or DRS_Depend_+ at 1 year; Supplementary Table 5).

Compared to the TBI-MS Model Testing cohort, TRACK-TBI participants were more likely to be white (81% vs 64%, p = 0.001), and appeared more severely injured (Table 2). Additionally, as expected, TRACK-TBI subjects were enrolled at an earlier post-injury timepoint (14 vs 24 days, p < 0.0001), were less likely to be discharged to inpatient rehabilitation (39% vs 100%, p < 0.0001), and were more likely to be dead or dependent (57% vs 26%, p < 0.0001) at 1-year post-injury (Table 2). These differences persisted even when analyses were restricted to participants in TBI-MS assessed at 14±3 days post-injury (Supplementary Table 6).

In the TRACK-TBI cohort, the Track-Aligned Model had an AUROC of 0.66 [0.53, 0.79] (p = 0.03) for predicting the primary outcome at 1 year. At the optimal cut-point established in the TBI-MS Testing Sample, the sensitivity was 83%, specificity was 39%, positive predictive value was 61%, and negative predictive value was 67%. The discrimination performance of the Track-Aligned Model did not differ from that of the IMPACT_core+CT_ model (AUROC 0.68 [0.55, 0.81]; 95% CI for AUROC difference: -0.2 to 0.2, p = 0.8).

## DISCUSSION

We used two large prospective studies to develop and characterize a dependency prediction model for patients with DoC after TBI. Within TBI-MS, our Discovery Sample, we found that a relatively small combination of variables (age, IVH, followed commands within 5 days of injury, and severity of functional and motor impairment at admission to rehabilitation) produces a well-calibrated estimate of the probability of functional dependency at 1-year. Roughly half of participants predicted to be dependent were dependent at 1-year post-injury; over 90% of those predicted not to be dependent were identified correctly. This finding was consistent regardless of whether deaths were included in the primary outcome. Though our model’s performance was diminished in an independent cohort (TRACK-TBI), it remained significantly better than chance and equivalent to the IMPACT prognostic model. This is a promising start for outcome prediction in patients with prolonged DoC after TBI.

Consistent with previous observations^25, 39^, despite severe injuries, a substantial proportion of patients did not meet criteria for dependency at 1-year post-injury. However, dependency, as defined here^4^, does not indicate complete independence. For instance, the definition of DRS_Depend_+ does not include patients with a DRS_Function_ item score of 3, which indicates a requirement for moderate assistance in the home.

The strongest independent predictors were advanced age and variables reflecting the severity of neurological impairment at rehabilitation admission. Patients with severe motor impairment (i.e., best motor exam of stereotyped movements such as extension) were nearly four-times as likely to be dependent as those without such impairment. The presence of IVH on initial CT scan was a weak but significant independent predictor, likely serving as a marker of diffuse axonal injury^40^.

In both datasets (TBI-MS and TRACK-TBI), our predictive model was more accurate in identifying patients who were *unlikely* to remain dependent than those *likely* to remain dependent. While it is inherently easier to develop a “rule-out” test for an uncommon outcome, this finding likely indicates that patients with signs of early neurological improvement, (e.g. command following within 5 days or localizing to noxious stimuli on the motor exam by admission to rehabilitation), have either preserved or recovered the substrate for functional recovery, which is consistent with prior work^41^ The absence of such signs, however, does not invariably lead to dependency at 1 year. For such patients, a more detailed assessment of the underlying neuroanatomical injury is needed, perhaps with advanced neuroimaging^42^ or electrophysiologic^43^ techniques.

Our findings suggest that the timing of the behavioral assessment is an important consideration for a model predicting outcomes after TBI. In patients with DoC at rehabilitation admission, GCS scores on hospital admission were not predictive of dependency, even in a univariate context. While TBI-MS does not collect all established predictors of outcome (e.g., pupillary reactivity at hospital admission), this finding generally suggests that among patients who survive with DoC on admission to rehabilitation, neurological impairment on hospital admission is not strongly related to recovery potential. However, among *all* patients with severe TBI, neurological impairment on admission is likely still a robust predictor of outcome.

Consistent with prior studies^44, 45^, in TBI-MS, the association between severe motor impairment and outcome was significantly stronger at 1-month compared to 2-weeks post-injury. In our TRACK-TBI DoC cohort, where all patients were assessed at 2 weeks post-injury, there was no clear association between severe motor impairment and outcome. More work is required to determine the optimal post-injury prognostication time, balancing the desire to provide an assessment as early as possible with the increased precision that likely comes from waiting.

Several factors likely contributed to the prediction model’s diminished performance in the external TRACK-TBI dataset. First, within TRACK-TBI, significantly fewer dependent than not-dependent outcome participants attended inpatient rehabilitation (16% vs 63%, respectively). Differences in the post-hospital environment and support systems that surround patients may alter the probability of achieving favorable functional outcomes. Second, missing outcomes and exposure variables in this TRACK-TBI cohort resulted in an effective sample size of fewer than 80 patients. Recent work establishing best practices for clinical prediction studies report unstable accuracy estimates in samples with fewer than several hundred patients^46^. Third, participants in TRACK-TBI appeared more severely injured than those in TBI-MS, even accounting for different first assessment times. Indeed, participants in TRACK-TBI had more than two times the incidence of dependency as those in TBI-MS. It is possible that predictor variables identified in TBI-MS may be less relevant in a population that is more severely injured. Finally, the TRACK-aligned model did not include the non-cortical contusion variable, which was not collected in TRACK-TBI. It is possible that this injury location variable would have improved model performance.

### Limitations

This work has several limitations. First, some important predictors may not have been identified due to excessive missingness. Second, our primary outcome used a relatively stringent dependency definition. Slightly different predictors may have been identified using less stringent outcome criteria. Third, while this represents the largest sample of patients with DoC after TBI studied to date, it is possible that additional associations would have been identified with an order of magnitude increase in sample size. Fourth, some patients included in the study because they were unable to follow commands may have had an isolated disturbance in language comprehension, which is difficult to dissociate from DoC^47^. However, this is unlikely to have occurred frequently, because isolated aphasia is uncommon after TBI^48^, and patients generally had multi-domain impairment, evidenced by high total DRS scores at rehabilitation admission. Finally, we used the IMPACT score here to benchmark our model’s performance. This comparison inherently disadvantages the IMPACT score because we are using a different outcome measure, different outcome time-point, and a different patient population from the sample in which IMPACT was initially developed^18^.

## Conclusions

We developed a 1-year functional dependency prediction model for patients with DoC after TBI in a large national TBI database (TBI-MS) and tested its external validity in an independent dataset (TRACK-TBI). The model incorporates a simple combination of variables, including age, severity of neurological impairment at time of assessment, command following in the first 5 days post-injury, and intraventricular hemorrhage. The model may serve as a reliable dependency rule-out tool for patients admitted to inpatient rehabilitation but has worse performance when applied to patients still receiving acute care. Although these findings illustrate the general challenges of developing a clinical prognostic model for a heterogeneous disease, this work represents an important step towards a systematic approach to predicting recovery for patients with DoC after TBI.

## Data Availability

All data produced in the present study are available upon reasonable request to the authors

## Acknowledgements

The contents of this manuscript were developed under a grant from the National Institute of Neurological Disorders and Stroke (grant number: U01NS086090) and the National Institute on Disability, Independent Living, and Rehabilitation Research (NIDILRR grant numbers 90DPCP0008-01-00, 90DP0039, 90DPTB0002 [Spaulding Rehabilitation Hospital and Indiana University). NIDILRR is a Center within the Administration for Community Living (ACL), Department of Health and Human Services (HHS). The contents of this manuscript do not necessarily represent the policy of NIDILRR, ACL, HHS, and you should not assume endorsement by the Federal Government.

