## Supplementary Material for "Predicting Functional Dependency in Patients with Disorders of Consciousness: A TBI-Model Systems and TRACK-TBI Study"

### SUPPLEMETARY MATERIAL

**Affiliations**
1 Division of Neurocritical Care, Department of Neurology, Brigham and Women's Hospital, Boston, MA, USA
2 Harvard Medical School, Boston, MA, USA
3 Department of Neurological Surgery, University of Washington, Seattle, Washington, USA.
4 Department of Biostatistics, University of Washington, Seattle, Washington, USA.
5 Center for Neurotechnology and Neurorecovery and Department of Neurology, Massachusetts General Hospital, Boston, MA, USA
6 Athinoula A. Martinos Center for Biomedical Imaging, Massachusetts General Hospital, Charlestown, MA, USA
7 Department of Physical Medicine and Rehabilitation, Spaulding Rehabilitation Hospital, Boston, MA USA
8 Department of Physical Medicine and Rehabilitation, Indiana University School of Medicine, Indianapolis, IN, USA
9 Departments of Neurosurgery and Neurology, University of Colorado School of Medicine, Aurora CO, USA
10 Department of Neurological Surgery, UCSF, San Francisco, CA USA
11 Brain and Spinal Cord Injury Center, Zuckerberg San Francisco General Hospital and Trauma Center, San Francisco, CA USA

Supp. Table 6: Comparison of Subjects with Comparable Lengths of Stay in TBI-MS …………..8
and TRACK-TBI
Supp. Figure 1: Distribution of Acute Care Lengths of Stay……………………………………....9
Supp. Figure 2: Track-TBI (Validation) Cohort Flowchart………………………………….....10
Supp. Figure 3: Relative Propensity Model Influence of TBI-MS Covariates…………………....11
Supp. Figure 4: Exposure Variable Collinearity ……………………..…………………………12
Supp. Figure 5: Exposure Variable Missingness……………………..…………………..…13
Supp. Figure 6: Performance of "Track-Aligned" Model in Testing Sample…………………..…14
Supp. Figure 7: Association Between $DRS_{Motor}$ and Primary Outcome Varies based on Days to First Post-Injury Assessment ………………………………………………………………………15
Supp. Figure 8: Dependency *Only* Prediction Model…………………..……………………..…16
Variable Definitions……………………………………………………………………...17-22

| **Supplementary Table 1:** TBI-MS Derivation vs Testing Cohorts | | | |
|---|---|---|---|
| | **Model Derivation (N = 1226)** | **Model Testing (N = 306)** | **P value** |
| ***DEMOGRAPHICS*** | | | |
| Age; Mean (SD) | 40 (19) | 42 (20) | 0.1 |
| Sex Male; N (%) | 946 (77) | 224 (73) | 0.1 |
| Race White; N (%) | 864 (71) | 197 (64) | 0.05 |
| Marital status; N (%) | | | |
| Single | 607 (50) | 140 (46) | 0.07 |
| Married | 388 (32) | 117 (38) | |
| Other | 230 (19) | 48 (16) | |
| Education; N (%) | | | |
| < HS | 341 (28) | 94 (31) | |
| HS diploma/GED | 324 (26) | 78 (26) | 0.8 |
| Some college | 190 (16) | 49 (16) | |
| Batchelor's Degree | 219 (18) | 51 (17) | |
| Employment; N (%) | | | |
| Competitively Employed | 647 (53) | 153 (5) | 0.1 |
| Student | 79 (6) | 13 (4) | |
| Unemployed | 343 (28) | 99 (32) | |
| Housing; N (%) | | | |
| Private Home | 1201 (98) | 299 (98) | 0.8 |
| Other | 23 (2) | 7 (2) | |
| ***CLINICAL CHARACTERISTICS*** | | | |
| Injury Mechanism; N (%) | | | |
| High-Velocity | 707 (58) | 161 (53) | 0.2 |
| Fall-related | 292 (24) | 87 (28) | |
| Low-Velocity/Other | 223 (18) | 57 (19) | |
| Injury Year; Mean (SD) | 2010 (6) | 2010 (6) | 0.7 |
| ED $GCS_{Verbal}$ < 3; N (%)[a] | 332 (27) | 84 (28) | 1 |
| ED $GCS_{Motor}$ < 5; N (%)[b] | 574 (47) | 123 (40) | 0.04 |
| ED $GCS_{Eye}$ < 2; N (%)[b] | 557 (45) | 118 (39) | 0.03 |
| Intubated; N (%) | 673 (55) | 151 (49) | 0.09 |
| Craniectomy; N (%) | 180 (18) | 47 (19) | 0.8 |
| CT Compression; N (%) | | | |
| None | 554 (55) | 137 (55) | |
| Cisterns present/MLS<5mm | 108 (11) | 23 (9) | 0.8 |
| Cisterns absent/MLS>5mm | 353 (35) | 90 (36) | |
| Intracranial Hypertension; N (%) | | | |
| No ICP spikes or unmonitored | 700 (57) | 182 (60) | |
| ICP ≥ 20 on one day | 184 (15) | 47 (15) | 0.7 |
| ICP ≥ 20 repeatedly or sustained | 322 (26) | 73 (24) | |
| SDH/SAH; N (%) | 1023 (88) | 241 (82) | 0.009 |
| EDH; N (%) | 137 (12) | 26 (9) | 0.2 |
| IVH; N (%) | 473 (41) | 127 (43) | 0.5 |
| Contusions | | | |
| Frontal; N (%) | 727 (59) | 182 (60) | 0.9 |
| Temporal; N (%) | 554 (45) | 127 (42) | 0.2 |
| Parietal; N (%) | 266 (22) | 63 (21) | 0.7 |
| Occipital; N (%) | 106 (9) | 20 (7) | 0.3 |
| Non-cortical; N (%) | 364 (30) | 102 (33) | 0.3 |
| Total #; Mean (SD) | 2 (2) | 2 (2) | 0.3 |

| | | | |
|---|---|---|---|
| Days post injury *[first assessment]**; Mean (SD) | 25 (12) | 24 (11) | 0.2 |
| DRS *[first assessment]*; Median (IQR)<br>Total Score<br>$DRS_{Function}$<br>$DRS_{Eye}$<br>$DRS_{Motor}$ | <br>22 (4)<br>5 (0)<br>0 (1)<br>1 (1) | <br>22 (4)<br>5 (0)<br>0 (1)<br>1 (1) | <br>0.4<br>0.6<br>0.4<br>0.7 |
| Rehab LOS; Mean (SD) | 49 (46) | 51 (45) | 0.6 |
| ***OUTCOMES*** | | | |
| DRS *[1-year]*; Median (IQR) | 5 (7) | 5 (7) | 0.2 |
| GOSE *[1-year]*; Median (IQR) | 4 (3) | 4 (3) | 0.5 |
| Dead or DRS-Depend +; N (%) | 325 (27) | 81 (27) | 1 |

*a* = in patient who is not intubated
*b* = in patient not receiving sedation/paralytic
*First assessment occurs at admission to rehabilitation
Abbreviations: SD = Standard Deviation; IQR = Interquartile Range; GED = General Educational Development Test; MLS = Midline Shift; SDH = Subdural Hemorrhage; SAH = Subarachnoid Hemorrhage; EDH = Epidural Hemorrhage; IVH = Intraventricular Hemorrhage; DRS = Disability Rating Scale; GOSE = Glasgow Outcome Scale Extended

| **Supplementary Table 2:** TBI-MS Univariate Association with Dependency ($p \leq 0.1$) | | |
|---|---|---|
| **Variable** | **Univariate OR** | **P value** |
| **DRS *[first assessment]**** | | |
| *Eye sub-score*<br>No eye opening<br>Eyes open with nox stim | 4.8 [2.8, 8.3]<br>1.9 [1.3, 2.8] | < 0.0001<br>0.0007 |
| *Total score category*<br>Extreme VS<br>VS | 4.4 [3.1, 6.3]<br>1.7 [1.3, 2.3] | < 0.0001<br>0.0003 |
| *Motor sub-score*<br>Flex/extend/no movement<br>Withdrawal | 3.5 [2.5, 4.8]<br>1.9 [1.4, 2.7] | < 0.0001<br>< 0.0001 |
| *Function sub-score*<br>Totally dependent: 24 hr care | 2.8 [1.9, 4.1] | < 0.0001 |
| Toilet sub-score<br>No awareness of continence | 2.2 [1.6, 3.2] | < 0.0001 |
| **Demographics** | | |
| Age ≥ 55 | 3.0 [2.2, 3.9] | < 0.0001 |
| Pre-Injury Employment:<br>Unemployed vs employed | 2.2 [1.7, 2.9] | < 0.0001 |
| Housing Status:<br>No private home vs private home | 2.1 [0.9, 4.6] | 0.07 |
| Pre-Injury Education:<br>HS Diploma vs < HS | 1.5 [1.1, 2.1] | 0.02 |
| **Clinical Characteristics** | | |
| IVH | 1.8 [1.4, 2.4] | < 0.0001 |
| SDH or SAH | 2.0 [1.2, 3.2] | 0.008 |
| Severe brain swelling:<br>Cisterns absent or MLS≥5mm | 1.7 [1.2, 2.2] | 0.0008 |
| Non-Cortical Contusion | 1.6 [1.2, 2.1] | 0.002 |
| Injury Mechanism:<br>Fall vs Low-Velocity | 1.5 [1.1, 2.2] | 0.02 |
| Parietal Contusion | 1.3 [1.0, 1.8] | 0.08 |
| Craniectomy | 1.3 [1.0, 1.9] | 0.08 |
| Sustained Intracranial Hypertension | 1.4 [1.0, 1.8] | 0.04 |
| Total Contusion # | 1.1 [1.0, 1.2] | 0.01 |
| Injury Mechanism:<br>High Velocity vs Low-Velocity | 0.7 [0.5, 1.0] | 0.04 |
| Followed cmds [within 5 days of injury] | 0.6 [0.4, 0.8] | 0.001 |

*First assessment occurs at admission to rehabilitation
Abbreviations: DRS = Disability Rating Scale; Flex = flexion; IVH = intraventricular hemorrhage; cmds = commands; HS = High School; nox = noxious; hr = hour; VS = Vegetative State

| **Supplementary Table 3: TBI-MS** Multivariate Dependency Prediction Model | | |
|---|---|---|
| **Variable** | **aOR** | **P value** |
| Age ≥ 55 | 4.3 [3.0, 6.1] | < 0.0001 |
| $DRS_{Motor}$ *[first assessment]*:* | | |
| *Flexion/extension/no movement* | 3.4 [2.4, 4.9] | < 0.0001 |
| *Withdrawal* | 1.7 [1.2, 2.4] | 0.002 |
| $DRS_{Function}$ *[first assessment]*: | | |
| *Totally dependent: 24 hr care* | 2.6 [1.7, 4.0] | < 0.0001 |
| Pre-Injury Employment: | | |
| *Unemployed* | 2.1 [1.5, 2.9] | < 0.0001 |
| *Student* | 0.7 [0.4, 1.2] | 0.2 |
| Underwent Craniectomy | 1.8 [1.3, 2.5] | 0.001 |
| IVH | 1.6 [1.2, 2.1] | 0.002 |
| Non-Cortical Contusion | 1.7 [1.2, 2.4] | 0.002 |
| Followed cmds [within 5 days of injury] | 0.5 [0.3, 0.7] | 0.0003 |

*First assessment occurs at admission to rehabilitation
Abbreviations: DRS: Disability Rating Scale; IVH: intraventricular hemorrhage; cmds: commands; hr: hour

| **Supplementary Table 4:** "TRACK-Aligned" Dependency Prediction Model | | |
|---|---|---|
| **Variable** | **aOR** | **P value** |
| Age ≥ 55 | 5.0 [3.6, 7.0] | < 0.0001 |
| $DRS_{Motor}$ *[first assessment]*:* | | |
| *Flexion/extension/no movement* | 3.5 [2.5, 5.0] | < 0.0001 |
| *Withdrawal* | 1.7 [1.2, 2.4] | 0.003 |
| $DRS_{Function}$ *[first assessment]*: | | |
| *Totally dependent: 24 hr care* | 2.5 [1.7, 3.8] | < 0.0001 |
| IVH | 1.7 [1.3, 2.2] | 0.0005 |
| Followed cmds [within 5 days of injury] | 0.5 [0.3, 0.7] | 0.0003 |

*First assessment occurs 2-weeks post injury

Abbreviations: DRS: Disability Rating Scale; IVH: intraventricular hemorrhage; cmds: commands; hr: hour

| **Supplementary Table 5:** TRACK-TBI External Validation Cohort Characteristics | | | | | |
|---|---|---|---|---|---|
| | | **1-Year Outcome** | | | |
| | **All (N = 124)** | $DRS_{Depend}$ - (N = 43) | $DRS_{Depend}$ + (N = 35) | Died (N = 22) | Missing (N = 24) |
| ***DEMOGRAPHICS*** | | | | | |
| Age; Mean (SD) | 40.3 (16.4) | 35.2 (16.3) | 40.6 (16.6) | 48.7 (15.4) | 41.4 (14.4) |
| Sex Male; N (%) | 95 (77) | 34 (79) | 23 (66) | 20 (91) | 18 (75) |
| Race White; N (%) | 100 (81) | 37 (86) | 26 (74) | 20 (91) | 17 (74) |
| Education Years; Mean (SD) | 12.4 (2.4) | 12.8 (2.6) | 11.7 (1.8) | 12.1 (2.9) | 12.9 (2.3) |
| ***CLINICAL CHARACTERISTICS*** | | | | | |
| Craniectomy; N (%) | 52 (43) | 18 (42) | 18 (51) | 11 (50) | 5 (23) |
| IVH; N (%) | 28 (24) | 9 (22) | 8 (25) | 4 (19) | 7 (32) |
| $DRS_{Function}$ *[first assessment]**; N (%) | | | | | |
| Not Totally-dependent [score = 1-4] | 1 (1) | 0 (0) | 0 (0) | 1 (5) | 0 (0) |
| Totally-dependent [5] | 122 (98) | 43 (100) | 34 (100) | 21 (95) | 24 (100) |
| $DRS_{Eye}$ *[first assessment]**; N (%) | | | | | |
| Opens eyes spont/to voice [0-1] | 35 (29) | 19 (44) | 10 (29) | 4 (18) | 2 (8) |
| Opens eyes to nox stim [2] | 32 (26) | 10 (23) | 9 (26) | 4 (18) | 9 (39) |
| Does not open eyes [3] | 56 (46) | 14 (33) | 16 (46) | 14 (64) | 12 (52) |
| $DRS_{Motor}$ *[first assessment]**; N (%) | | | | | |
| Localizing [1] | 13 (11) | 5 (12) | 6 (17) | 0 (0) | 2 (9) |
| Withdrawal [2] | 34 (28) | 13 (30) | 8 (23) | 3 (14) | 10 (43) |
| Flex/extend/no mvmt [3-5] | 76 (61) | 25 (59) | 21 (60) | 19(86) | 11 (47) |
| Days post injury *[first assessment]**; Mean (SD) | 14.4 (2.5) | 14.0 (2.7) | 14.7 (2.3) | 14.1 (2.5) | 15.1 (2.5) |
| Followed cmds first 5 days; N (%) | 20 (20) | 11 (30) | 5 (20) | 1 (5) | 3 (15) |
| $IMPACT_{Core}$ score; Mean (SD) | 43 (19) | 40 (21) | 45 (15) | 49 (19) | 41 (18) |
| $IMPACT_{Core+CT}$ score; Mean (SD) | 45 (20) | 40 (21) | 49 (18) | 57 (18) | 40 (17) |
| Discharged to Rehab; N (%) | 45 (39) | 27 (63) | 8 (23) | 1 (7) | 9 (39) |
| ***OUTCOMES*** | | | | | |
| 12-Month GOSE; Mean (SD) | 3.5 (2.0) | 5.1 (1.7) | 3.1 (0.4) | -- | -- |

*First assessment occurs 2-weeks post injury

Abbreviations: SD standard deviation; IVH intraventricular hemorrhage; nox noxious; DRS Disability Rating Scale; cmds commands; GOSE Glasgow Outcome Scale Extended

| **Supplementary Table 6:** Comparison of Subjects with Comparable Lengths of Stay in TBI-MS and TRACK-TBI | | | |
|---|---|---|---|
| | **TBI-MS Cohort LOS 11-17 days (N = 380)** | **TRACK-TBI External Validation Cohort (N = 124)** | **P value** |
| ***DEMOGRAPHICS*** | | | |
| Age; Mean (SD) | 45 (21) | 40 (16) | 0.008 |
| Sex Male; N (%) | 273 (72) | 95 (77) | 0.4 |
| Race White; N (%) | 253 (67) | 100 (81) | 0.004 |
| Education Years; Mean (SD) | 12 (3) | 12 (2) | 0.8 |
| ***CLINICAL CHARACTERISTICS*** | | | |
| Craniectomy; N (%) | 38 (12) | 52 (43) | < 0.0001 |
| IVH; N (%) | 122 (34) | 28 (24) | 0.02 |
| $DRS_{Function}$ *[first assessment]**; N (%)<br>Not Totally-dependent [score = 1-4]<br>Totally-dependent [5] | <br>128 (34)<br>252 (66) | <br>2 (2)<br>122 (98) | < 0.0001 |
| $DRS_{Eye}$ *[first assessment]**; N (%)<br>Opens eyes spont/to voice [0-1]<br>Opens eyes to nox stim [2]<br>Does not open eyes [3] | <br>340 (89)<br>24 (6)<br>16 (4) | <br>35 (28)<br>32 (26)<br>56 (45) | < 0.0001 |
| $DRS_{Motor}$ *[first assessment]**; N (%)<br>Localizing [1]<br>Withdrawal [2]<br>Flex/extend/no mvmt [3-5] | <br>283 (74)<br>58 (15)<br>39 (10) | <br>13 (10)<br>34 (28)<br>76 (62) | < 0.0001 |
| Days post injury *[first assessment]**; Mean (SD) | 14 (2) | 14 (3) | 1 |
| Followed cmds first 5 days; N (%) | 155 (41) | 20 (20) | 0.0002 |
| Discharged to inpatient rehab; N (%) | 380 (100) | 45 (39) | < 0.0001 |
| ***OUTCOMES*** | | | |
| $DRS_{Depend}$ + *[1-year]*; N (%) | 30 (10) | 35 (35) | < 0.0001 |
| Died; N (%) | 21 (7) | 22 (22) | < 0.0001 |
| GOSE 2-3 *[1-year];* N (%) | 65 (22) | 39 (39) | 0.002 |
| Missing Outcome; N (%) | 88 | 24 | 0.4 |

*****First assessment occurs 2-weeks post injury
Abbreviations: LOS Length of Acute Care Stay; SD standard deviation; IVH intraventricular hemorrhage; nox noxious; DRS Disability Rating Scale; spont spontaneous; cmds commands; GOSE Glasgow Outcome Scale Extended (GOSE)

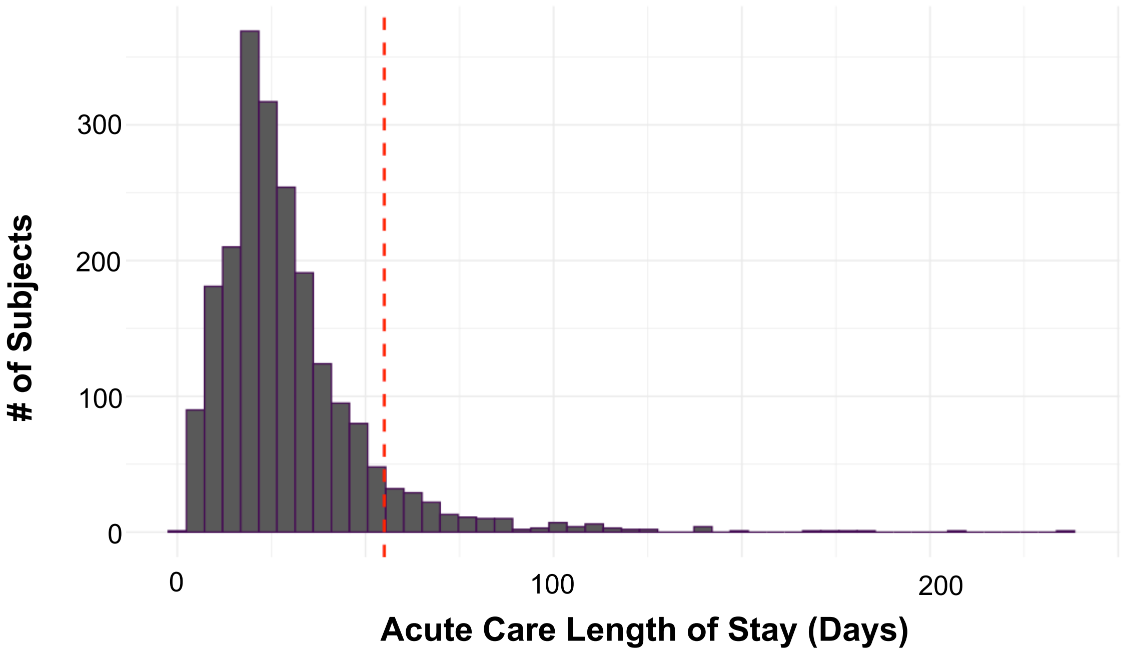

**Supplementary Figure 1: Distribution of Acute Care Lengths of Stay**
Histogram of acute care lengths of stay. Red dashed line indicates 1.5 x Interquartile Range, beyond which subjects were excluded from the study.

**TRACK-TBI (EXTERNAL VALIDATION) COHORT**

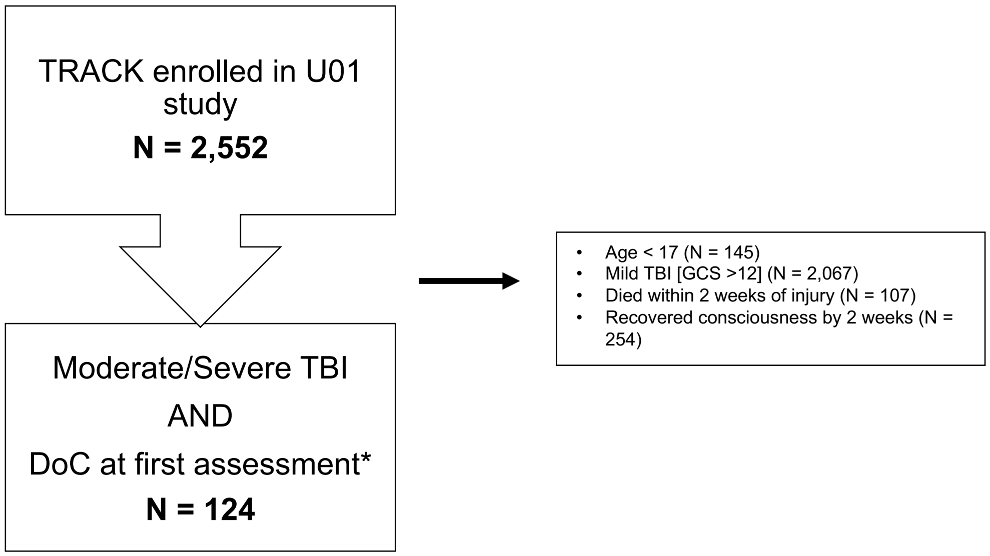

**Supplementary Figure 2: Track-TBI (Validation) Cohort Flowchart**
TRACK-TBI (Validation Cohort) CONSORT flow diagram.
*first assessment occurred at 2-weeks post injury
Abbreviations: DoC: disorders of consciousness; GCS: Glasgow Come Scale; TBI: traumatic brain injury

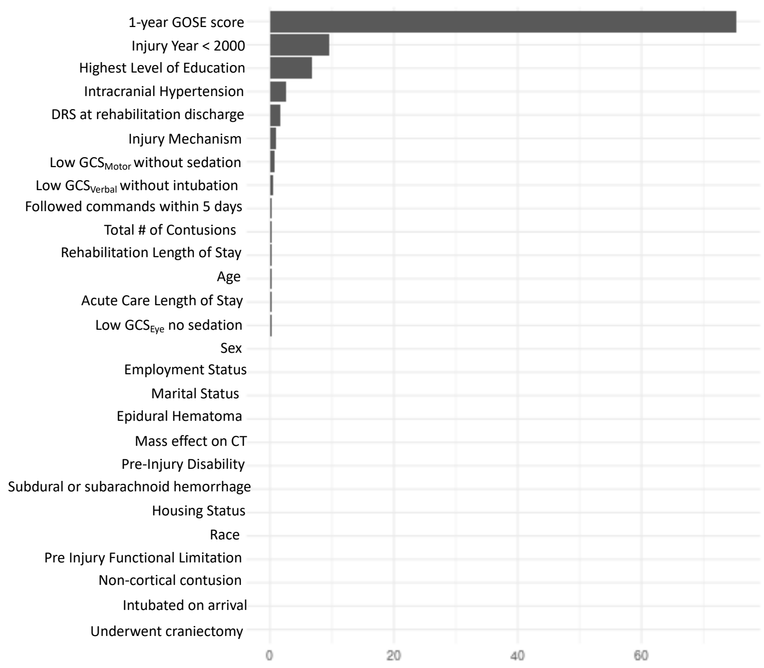

**Supplementary Figure 3: Relative Propensity Model Influence of TBI-MS Covariates**
All variables screened for the gradient boosted inverse propensity model. Size of each bar indicates relative contribution to the propensity score.

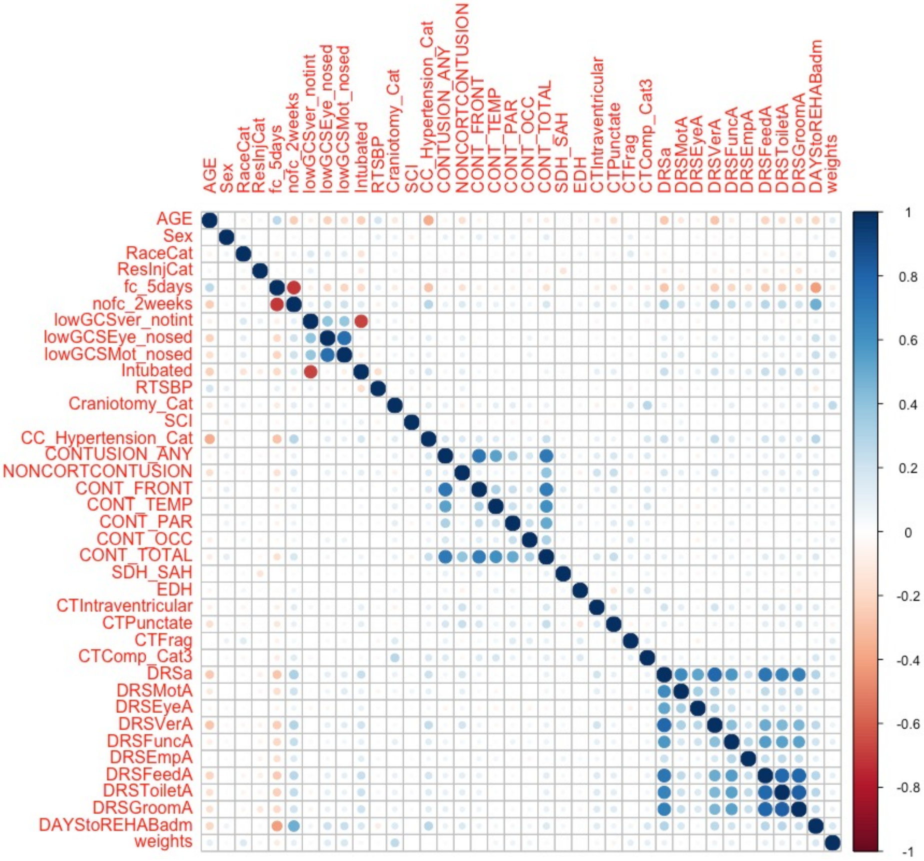

**Among ≥ 0.7 colinear pairs below, red variable excluded from further analyses:**

[LowGCSEyeNoSed vs lowGCSMotnosed ]
[no_cmds_2weeks vs flw_cmds_5days]
[ContusionAny vs ContusionFrontal ]
[ContusionTotal or ContusionANY]
[DRSa (Total Score) vs DRSVerA]
[DRSa (Total Score) vs DRSFeedA]
[DRSToiletA vs DRSFeedA]
[DRSToiletA vs DRSGroomA]

**Supplementary Figure 4: Exposure Variable Collinearity**

Correlation matrix (Spearman's Rho) for all exposure variables. Among variable pairs with Spearman's Rho ≥ 0.7 (listed to the right of the plot) one variable was excluded (red) from further analysis. Abbreviations: DRS = Disability Rating Scale; LowGCSEyeNoSed = $GCS_{Eye}$ < 2 and patient not sedated; LowGCSMotnosed = $GCS_{Motor}$ ≤ 4 and patient not sedated/paralyzed; no_cmds_2weeks = no command following within 2 weeks of injury; flw_cmds_5days = command following within 5 days of injury

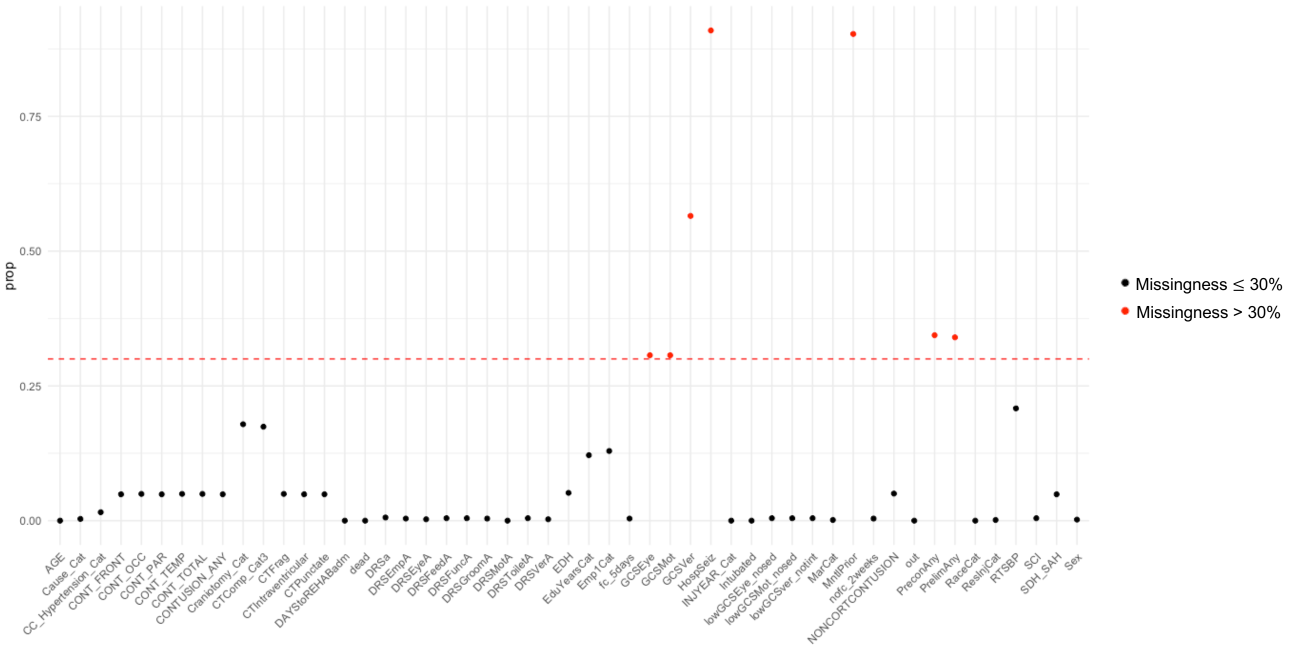

**Supplementary Figure 5: Exposure Variable Missingness**
The proportion of missing data for each candidate exposure variable. Variables with > 30% missing data (red) were excludm further analysis.

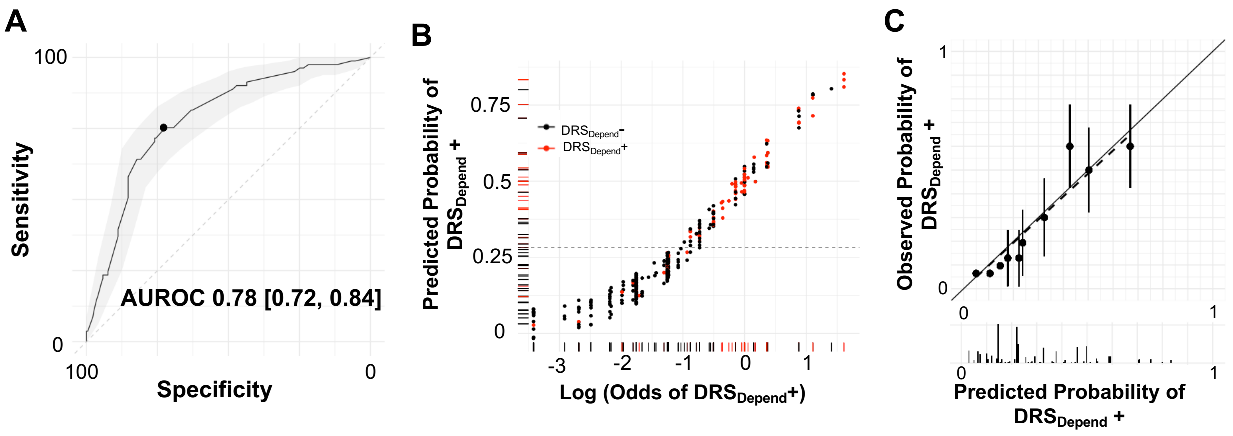

**Supplementary Figure 6: Performance of "Track-Aligned" Model in Testing Sample**
(A) Area under the Receiver Operating Characteristic Curve for Track-Aligned Dependency prediction model within the TBI-MS Testing Sample. Shaded area reflects the 95% confidence interval. (B) Rug plot showing predicted dependency probabilities (y axis) and the logarithm of the odds of dependency (x axis). Each dot represents one subject and each subject is also entered as a tick on each axis. Color indicates 1-year outcome (red: $DRS_{Depend}$+, black: $DRS_{Depend}$-). The dashed line represents the Youden J cut-point derived from (A). $DRS_{Depend}$+ subjects cluster towards the upper right quadrant of the plot. (C) Calibration plot showing predicted dependency probabilities (x axis) and observed probabilities (y axis). Each dot represents a 10-percentile bin of predicted probabilities. Error bars represent 95% confidence intervals. Histogram below the x axis shows the relative density of observations in each predicted probability bucket.

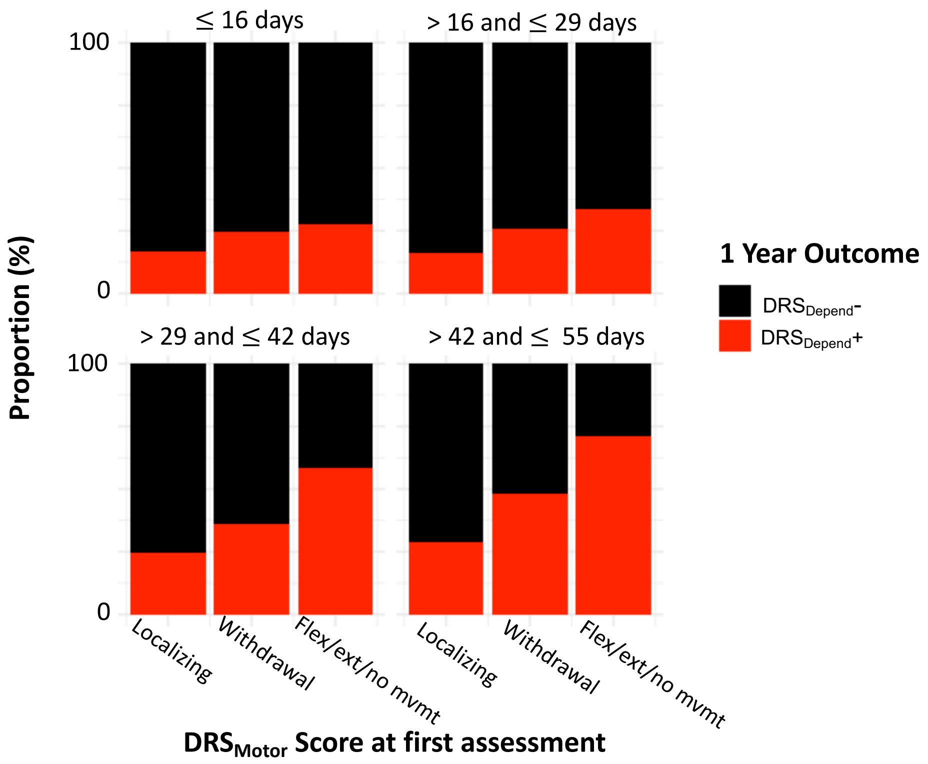

**Supplementary Figure 7: Association Between $DRS_{Motor}$ and Primary Outcome Varies based on Days to First Post-Injury Assessment.**

In the TBI-MS Model Derivation cohort, the proportion of each 1-year outcome is shown based on $DRS_{Motor}$ scores at time of first assessment (rehabilitation admission). Each quadrant represents one strata of days from injury to rehabilitation admission. As days between injury and rehabilitation admission increase, the difference in the proportion of patients with death or $DRS_{Depend}$+ 1-year outcomes across $DRS_{Motor}$ scores becomes more pronounced. For example, for participants assessed for the first time (i.e., admitted to rehabilitation) within approximately 2 weeks post-injury, the $DRS_{Motor}$ item score is not strongly associated with outcome (height of top left panel red bars are similar). However, for participants assessed for the first time (i.e., admitted to rehabilitation) 42-55 days post-injury, there is a much clearer association between more impaired $DRS_{Motor}$ item scores and 1-year dependency ($DRS_{Depend}$+). Tested statistically, there was a significant interaction (in the full model) between $DRS_{Motor}$ score of Flexion/extension/no movement and time to first assessment, with each additional post injury day increasing $DRS_{Motor}$ aOR by 1.04 [1.01, 1.08] (p=0.01). Thus, a more impaired motor exam later post-injury may be more predictive of outcome than a more impaired motor exam early in recovery. Abbreviations: Flex: flexion, Ext: extension, Mvmt: movement

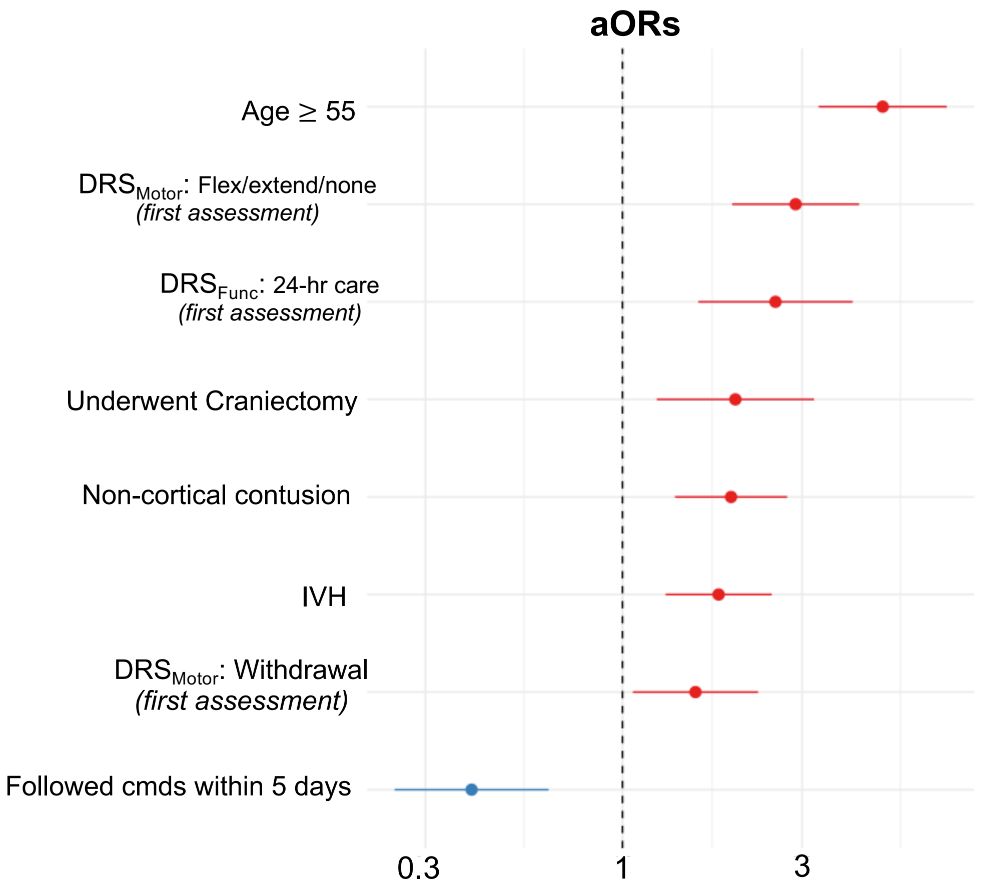

**Supplementary Figure 8: Dependency *Only* Prediction Model**
We derived new inverse propensity weights and fit a new *dependency-only* prediction model in the TBI-MS Discovery Sample (N=1429; Derivation: N=1145, Testing: N=284). The resulting model was nearly identical to our initial model, with AUROC of 0.78 [0.71, 0.84], sensitivity 68%, specificity 75%, PPV of 42% and NPV 90%. A forest plot of adjusted odds ratios (aORs) for variables included in the dependency-only prediction model (i.e., deaths excluded from analysis). Abbreviations: DRS: Disability Rating Scale, Cmds: commands, IVH: intraventricular hemorrhage, SDH: subdural hematoma, SAH: subarachnoid hemorrhage.

**Predicting Functional Dependency in Patients with Disorders of Consciousness after Traumatic Brain Injury: A TBI-Model Systems and TRACK-TBI Study**

Samuel B. Snider MD[1,2], Nancy R. Temkin PhD[3,4], Jason Barber MS[3], Brian L. Edlow MD[2,5,6], Joseph T. Giacino PhD[2,7], Flora M. Hammond MD[8], Saef Izzy MD[1,2], Robert G. Kowalski, MBBCh, MS[9], Amy J. Markowitz JD[10], Craig A. Rovito MD[2,7], Shirley L. Shih MD[2,7], Ross D. Zafonte DO[2,7], Geoffrey T. Manley MD PhD[10,11], Yelena G. Bodien PhD[2,5,7] and the TRACK-TBI investigators.

### <u>SUPPLEMETARY MATERIAL:</u> *Variable Definitions*

**Affiliations**
1 Division of Neurocritical Care, Department of Neurology, Brigham and Women's Hospital, Boston, MA, USA
2 Harvard Medical School, Boston, MA, USA
3 Department of Neurological Surgery, University of Washington, Seattle, Washington, USA.
4 Department of Biostatistics, University of Washington, Seattle, Washington, USA.
5 Center for Neurotechnology and Neurorecovery and Department of Neurology, Massachusetts General Hospital, Boston, MA, USA
6 Athinoula A. Martinos Center for Biomedical Imaging, Massachusetts General Hospital, Charlestown, MA, USA
7 Department of Physical Medicine and Rehabilitation, Spaulding Rehabilitation Hospital, Boston, MA USA
8 Department of Physical Medicine and Rehabilitation, Indiana University School of Medicine, Indianapolis, IN, USA
9 Departments of Neurosurgery and Neurology, University of Colorado School of Medicine, Aurora CO, USA
10 Department of Neurological Surgery, UCSF, San Francisco, CA USA
11 Brain and Spinal Cord Injury Center, Zuckerberg San Francisco General Hospital and Trauma Center, San Francisco, CA USA

| Exposure variables (TBI-MS) | | | | |
|---|---|---|---|---|
| **Demographics** | **Pre-Injury** | **Injury** | **Hospital** | **CT** |
| Age | Pre-injury disability | GCS motor | Followed commands within 5 days | Subdural or Subarachnoid Hemorrhage (SDH/SAH) |
| Sex | Pre-injury limitation | GCS Verbal | No commands for 14 days | Intraventricular Hemorrhage (IVH) |
| Race | Pre-Injury Marital status | GCS Eye | Underwent Craniectomy | Epidural Hemorrhage (EDH) |
| Year of Injury | Pre-Injury Residence | LowGCSEye, not sedated | Spinal cord injury | Punctate/Petechial hemorrhage |
| | Pre-Injury Education | LowGCSMotor, not sedated | Seizures | Intracranial Bone Fragment |
| | Pre-Injury Employment | LowGCSVerbal, not intubated | Intracranial hypertension | Any Contusion |
| | Pre-Injury Mental health problem | Intubated on arrival | Days post injury of initial DRS assessment | Non-Cortical Contusion |
| | | Systolic Blood Pressure on arrival | Initial DRS sub-items at admission to rehab (categorized as below) | Frontal/Temporal/Parietal/ Occipital Contusion |
| | | Spinal Cord Injury? | Initial DRS total score category on admission to rehab | CT Compression |
| | | Injury Mechanism | Days between Injury and 1-year follow-up | Total # of Contusions |

**VARIABLE DEFINITIONS PROVIDED BELOW:**

### <u>DEMOGRAPHICS</u>

**Age:**

Recoded from continuous to binary variable: < or ≥55

**Race:**

White
non-white

**Year of Injury:**

< 2000
≥ 2000

### PRE-INJURY

**Pre-Injury Disability:**

Absent
Present: *A pre-existing condition resulting in blindness, deafness, or substantial limitation in one or more basic physical activities such as walking, climbing stairs, reaching, lifting or carrying.*

**Pre-Injury Limitation:**

Absent
Present: *A pre-existing condition resulting in:*

*difficulties learning, remembering or concentrating*
*difficulties in dressing, bathing or getting around inside the home*
*difficulties going outside the home alone to shop or visit a doctor*
*difficulties working at a job or business*

**Pre-Injury Marital status:**

Single
Married
Other

**Pre-Injury Residence:**

Private home
All others: *Nursing home, adult home, correctional institution, hotel, homeless, hospital: acute care, hospital: rehabilitation, hospital: other*

**Pre-Injury Mental Health Problem** (in year preceding injury):

No
Yes

**Pre-Injury Education:**

< High School
High School diploma or GED
Some college
Batchelor's degree or beyond

**Pre-Injury Employment:**

Competitively employed
Student
Unemployed or not in labor force

### Clinical Characteristics

**LowGCSverbal, not intubated:**
***Synthetic variable created because GCSVerbal scored as missing (rather than 1) if intubated, sedated or paralyzed***

Absent:

Present*: (Patient not intubated/sedated/paralyzed and GCS Verbal score < 3)*

**LowGCSMotor, not sedated:**
***Synthetic variable created because GCSMotor scored as missing (rather than 1) if sedated or paralyzed***

Absent:

Present: *(Patient not sedated/paralyzed and GCS Motor score ≤ 4)*

**LowGCSEye, not sedated:**
***Synthetic variable created because GCSEye scored as missing (rather than 1) if sedated or paralyzed***

Absent:

Present: *(Patient not sedated/paralyzed and GCS Eye score < 2)*

**Systolic Blood Pressure:**

0 if in arrest on arrival

**Intracranial Hypertension:**

0: *unmonitored or no episodes of ICP* ≥ 20 mmHg

1: *single episode of ICP ≥ 20 mmHg during one 24-hour period*

2: *ICP ≥20 mmHg during multiple 24-hour periods or sustained elevation for one 24-hour period*

**Injury Mechanism:**

High velocity*: (motor vehicle, motorcycle, bicycle, All-Terrain Vehicle, All-Terrain Cycle, or other vehicular)*

Fall-related: *(any head injury related to a fall)*

Other/Low velocity: *(gunshot wound, assaults with blunt instrument, other violence, water sports, field/track sports, gymnastic activities, winter sports, air sports, other sports, hit by falling/flying object, pedestrian, or other/unclassified)*

**Followed commands within 5 days:**

Absent

Present: (*Abstracted from patients medical record: documentation, within 5 days of injury, of command following two times within a 24 hour period or documentation of* $GCS_{motor}$ *score of 6 two times within a 24 hour period)*

**No commands for 14 days:**

Absent:

Present: (*Abstracted from patients medical record: Absence of documentation, within 14 days of injury, of command following two times within a 24 hour period or documentation of* $GCS_{motor}$ *score of 6 two times within a 24 hour period)*

**First assessment $DRS_{Motor}$ Re-coded**

0: (*$DRS_{Motor}$ 1) Localizes noxious stimuli*
1: ($DRS_{Motor}$ 2) *Withdraws to noxious stimuli*
2: ($DRS_{Motor}$ 3-5) *Flexes, extends, or does not move to noxious stimuli*

**First assessment $DRS_{Eye}$ Re-coded**

0: (*$DRS_{Eye}$ 0-1)Eyes open spontaneously or with verbal stimulation*
1: (*$DRS_{Eye}$ 2) Eyes open with noxious stimulation*
2: (*$DRS_{Eye}$ 3) Eyes do not open*

**First assessment $DRS_{Verbal}$**

0: Oriented
1: Confused
2: Making any verbal utterances
3: No sounds or speech

**First assessment $DRS_{Groom}$**

0: *Complete awareness*
1: *Partial awareness*
2: *Minimal awareness*
3: No *awareness*

**First assessment $DRS_{Feed}$**

0: *Complete awareness*
1: *Partial awareness*
2: *Minimal awareness*
3: No *awareness*

**First assessment $DRS_{Toilet}$ Re-coded**

0: *($DRS_{Toilet}$ 0-2) Complete awareness to Minimal Awareness*
1: *($DRS_{Toilet}$ 3) No awareness*

**First assessment $DRS_{Functional}$ Re-coded**

0: (*$DRS_{Functional}$ 0-4) Completely independent to Markedly Dependent, assistance required*
*for all major activities*
1: (*$DRS_{Functional}$ 5) Totally dependent: 24 hour nursing care*

**First assessment $DRS_{Employment}$ Re-coded**

0: (*$DRS_{Employment}$ 0-2)*: *Not restricted to sheltered workshop, non-competitive*
1: (*$DRS_{Employment}$ 3)*: *Not employable*

**First assessment $DRS_{Total}$ Category Recode**

0-21 (None to Extremely Severe Disability)
22-24 (Vegetative State)
25-29 (Extreme Vegetative State)

### CT-SCAN

*(CODED BASED ON ALL CT REPORTS WITHIN 7 DAYS OF INJURY; MRI NOT INCLUDED)*

**CT Compression:**

None
Mild (*cisterns present, shift < 5mm)*
Severe *(Midline shift > 5mm or cisterns absent)*

**SDH/SAH:**

Absent
Present *(Subdural or subarachnoid hemorrhage present)*

**IVH:**

Absent
Present (Intraventricular hemorrhage present)

**EDH:**

Absent
Present (Epidural hemorrhage present)

**Intracranial Bone Fragment:**

Absent
Present

**Punctate/Petechial hemorrhage:**

Absent
Present

**Frontal/Temporal/Parietal/Occipital Contusion:**

Absent
Present *(left-sided, right-sided or side not specified)*

**Unknown-location Contusion:**

Absent
Present *(location not specified)*

**Non-cortical Contusion:**

Absent
Present *(left-sided, right-sided or side unspecified parenchymal but non-cortical contusion)*

**Any Contusion:**

Absent
Present *(Any frontal, temporal, parietal, occipital or location unspecified contusion. Non-cortical contusions not included).*

**Total Contusions:**

Total number of scored contusion locations in the brain (summed across left and right hemisphere)
